# Beyond spouses: primary social supporter selection among pregnant women enrolled in the Support-Moms study in southwestern Uganda – a cross-sectional study

**DOI:** 10.64898/2026.11.16.26368248

**Authors:** Esther C. Atukunda, Henry Ochola, Elly B. Atuhumuza, Jessica Haberer, Van T. Nghiem, Peter Waiswa, Micheal Kanyesigye, Leevan Tibaijuka, Charles Lugaaju, Justus Asasira, Stuart Turanzomwe, Celestino Obua, Mark J. Siedner, Lynn T. Matthews, Godfrey R. Mugyenyi

## Abstract

Social support during pregnancy can influence women’s engagement with antenatal care and other maternal health behaviours. Maternal health programmes in sub-Saharan Africa have often emphasised male partner involvement, although women may draw on broader social networks. Evidence on whom women identify as their primary supporter during pregnancy remains limited. We examined the prevalence and factors associated with non-spousal primary supporter selection among pregnant women in southwestern Uganda.

We conducted a cross-sectional analysis of baseline data from the Support-Moms study, a Type 1 hybrid effectiveness–implementation randomised controlled trial conducted in Mbarara City, Mbarara District, and Mitooma District, southwestern Uganda. Pregnant women enrolled between 27 February and 17 December 2024 identified two social supporters before randomisation; the first-nominated individual was classified as the primary supporter. The primary outcome was selection of a non-spousal primary supporter. Modified Poisson regression with robust variance was used to estimate crude and adjusted prevalence ratios (aPRs) with 95% confidence intervals (CIs).

Among 824 women, 223 (27.1%; 95% CI 24.1–30.3) selected a non-spousal primary supporter. Friends (30.5%) and other relatives (28.7%) were the most frequently nominated non-spousal supporters, followed by siblings (17.9%) and mothers (16.6%). Non-spousal selection was more prevalent among women recruited in Mbarara District (aPR 1.50, 95% CI 1.09–2.08) and Mitooma District (aPR 2.52, 95% CI 1.77–3.60) than Mbarara City, and among women who were separated, divorced, widowed, or never married (aPR 2.35, 95% CI 1.76–3.14), had an unplanned pregnancy (aPR 1.39, 95% CI 1.10–1.77), had a >10-year relationship with their nominated supporter (aPR 1.93, 95% CI 1.39–2.69), or were aged <20 years (aPR 1.48, 95% CI 1.01–2.15).

Although spouses were the primary supporters for most women, more than one-quarter nominated someone else, most commonly a friend or relative. These findings suggest that pregnancy support extends beyond marital relationships and may be shaped by relational status, social context, pregnancy intention, age, and established social ties. Antenatal care programmes may benefit from complementing partner-focused approaches with mechanisms to engage other support figures identified by women.

## Introduction

Pregnancy is a critical period during which social support plays an important role in shaping maternal health behaviours, emotional well-being, and engagement with antenatal care services [1–4]. Evidence from diverse settings indicates that supportive social environments during pregnancy are associated with improved maternal mental health, increased uptake of antenatal care services, and better maternal and neonatal outcomes [5–8]. Social support during pregnancy may take multiple forms, including emotional encouragement, financial assistance, informational guidance, and practical help with daily activities [9]. These forms of support often arise from a woman’s broader social network, including partners, family members, friends, and community members [9].

A social network perspective provides a useful framework for understanding pregnancy support. Social network theory conceptualises individuals as embedded within networks of relationships through which social resources and support can be accessed and exchanged [10, 11]. The structure and composition of these networks, as well as the functions provided by different social ties, may influence health-related behaviours and access to care [10, 11]. During pregnancy, these social ties may include spouses or partners, family members, friends, neighbours, and community members, whose roles and influence may vary according to women’s circumstances and the nature of their relationships [12].

In sub-Saharan Africa (SSA), maternal health programmes have increasingly emphasised male partner involvement as a key strategy for improving maternal and newborn health outcomes [13–17]. Over the past two decades, numerous interventions have sought to promote partner participation in antenatal care visits, encourage joint decision-making during pregnancy, and increase uptake of HIV testing and prevention of mother-to-child transmission services [14, 17–19]. Evidence suggests that male partner engagement can improve maternal health service utilisation, enhance adherence to prevention of mother-to-child transmission interventions, and strengthen household support for maternal health behaviours [13, 18, 20]. Consequently, many maternal health policies and programmes place considerable emphasis on male partners as sources of support during pregnancy [20].

However, emerging evidence suggests that pregnancy support systems are often embedded within broader kinship and social networks rather than exclusively within intimate partnerships. In many African settings, family members, particularly mothers, grandmothers, and other female relatives, play central roles in supporting pregnant women through emotional guidance, financial assistance, and practical caregiving support [21, 22]. Younger women and those experiencing unintended pregnancies may be particularly likely to rely on extended family or social networks for support, especially in contexts where partner relationships are unstable or non-cohabiting [15, 21–23]. These patterns suggest that pregnancy support may function within broader social ecosystems rather than solely within partner-centred relationships.

Despite the recognised importance of social support during pregnancy, fewer studies have specifically examined whom pregnant women themselves identify as their primary source of support. Much of the existing literature has focused on male partner involvement or women’s general perceptions and receipt of social support rather than on the specific individual women identify as their primary supporter [9, 12, 17]. Understanding who women nominate as their primary supporter may therefore provide valuable insights into the structure of maternal support networks and the social dynamics that influence pregnancy care.

To address this gap, we conducted a cross-sectional analysis of data from pregnant women participating in the Support-Moms study in southwestern Uganda [24]. Support-Moms is a Type 1 hybrid effectiveness-implementation randomised controlled trial evaluating the integration of a patient-centred mHealth social support intervention into routine antenatal care (ANC). At enrolment, before randomisation and exposure to the intervention, participants identified two individuals from their existing social networks who could provide support during pregnancy. These baseline supporter nominations provided an opportunity to examine, as a secondary cross-sectional analysis, whom women identified as their primary supporter and the factors associated with selecting a non-spousal supporter. Specifically, we aimed to determine the prevalence of non-spousal primary supporter selection during pregnancy and identify factors associated with this choice.

## Materials and Methods

### Study design and setting

This study was a cross-sectional analysis of baseline data collected in the Support-Moms study, a type 1 hybrid effectiveness-implementation RCT evaluating the integration of a patient-centred mHealth social support intervention into routine ANC. The protocol for this trial is published elsewhere [24].

The study was conducted across publicly funded health facilities in Mbarara City, Mbarara District, and Mitooma District in southwestern Uganda, representing predominantly urban, peri-urban, and rural settings, respectively. Participating facilities included a regional referral hospital and Health Centres IV and III. The study areas comprise predominantly subsistence-based livelihoods and face persistent gaps in maternal health service utilisation. For the present analysis, we used baseline data collected at enrolment before randomisation into the parent trial.

### Study population and recruitment

The source population comprised pregnant women enrolled in the Support-Moms study between 27 February and 17 December 2024. Women were eligible for the parent study if they: (1) were <20 weeks’ gestation and had not yet initiated antenatal care; (2) resided within the catchment area of a participating health facility; (3) were emancipated minors or adults aged ≥18 years; (4) had access to a mobile phone with network reception at home; (5) were able to identify at least two social supporters residing within the study districts; and (6) provided written informed consent [24]. For this secondary cross-sectional analysis, we included all women enrolled in the parent study who had complete data on their first-nominated social supporter relationship.

### Variables

### Outcome variable, data source, and measurement

The primary outcome was non-spousal primary social supporter selection, defined as nomination of a person other than the participant’s spouse as her first identified social supporter. The relationship to the first-nominated supporter was recorded using predefined relationship categories and subsequently classified for analysis as spousal or non-spousal. Non-spousal categories included friend, neighbour, mother, sibling, VHT/healthcare provider, and other relative.

At enrolment and before randomisation, participants were asked to identify two individuals from their existing social networks who could provide support during pregnancy. The first-nominated individual was designated the primary social supporter and the second-nominated individual the secondary social supporter. The present analysis focused on the primary social supporter because this was the participant’s first nomination.

Participants who did not nominate their spouse as their primary supporter were asked to include their spouse among their secondary supporters. Thus, the primary supporter nomination reflects whom women selected first within the supporter-nomination structure of the parent study, rather than an unconstrained ranking of all available social relationships.

### Explanatory variables, data source, and measurement

The explanatory variables were selected a priori based on existing literature, contextual relevance, and conceptual plausibility [10–12]. Variables were selected to capture sociodemographic, reproductive, clinical, relationship, and social characteristics that could plausibly influence the composition or mobilisation of women’s social support networks during pregnancy.

Sociodemographic characteristics included maternal age (<20, 20–34, and ≥35 years), parity, highest educational attainment, occupation (not employed, informal employment, or professional employment), household monthly income, recruitment site, and marital status. Maternal age, parity, education, occupation, and marital status were self-reported. Recruitment site was classified according to the participant’s recruitment site as Mbarara City (predominantly urban), Mbarara District (predominantly peri-urban), or Mitooma District (predominantly rural). Household monthly income was self-reported and summarised as a continuous variable for descriptive analyses and categorised as ≤150,000 UGX versus >150,000 UGX for regression analysis, consistent with income thresholds used in previous studies in Uganda [25, 26].

Reproductive and relationship characteristics included gestational age at recruitment (≤12 or ≥13 weeks), pregnancy intention and planning, duration of the relationship with the identified social supporter (<2 years, 2–5 years, 6–10 years, and >10 years), spouse–woman age difference (spouse younger or 0–4 years older, 5–9 years older, or ≥10 years older), and partner support of the current pregnancy. These characteristics were obtained through interviewer-administered questions at baseline.

Clinical and social characteristics included HIV status, history of gender-based violence, membership in a savings or support group, distance to the nearest health facility providing antenatal care, and household food insecurity. HIV status was self-reported. History of gender-based violence and relationship characteristics were assessed using questions adapted from validated tools used in Uganda [27]. Food insecurity was assessed using the Household Food Insecurity Access Scale (HFIAS) [28]. Distance to the nearest health facility providing antenatal care was self-reported.

Characteristics of the nominated social supporters, including sex, age, occupation, and relationship to the participant, were also recorded at baseline and were used to describe the composition of women’s social support networks.

All baseline data were collected before randomisation using structured interviewer-led questionnaires administered in Runyankole and Rukiga. Data were entered electronically using REDCap, a secure web-based data capture platform, to facilitate data completeness and quality control [29]. Further details regarding instrument development, data capture systems, and quality assurance procedures are described in the published study protocol [24].

### Study Sample size

This study was a secondary cross-sectional analysis of baseline data from the Support-Moms trial, in which 824 pregnant women were enrolled. The sample size was determined for the parent RCT; no additional sample size calculation was performed for this analysis.

To assess the precision of the prevalence estimate, we used the standard error of a proportion, SE = √[p(1−p)/n], where p is the estimated prevalence and n is the sample size. Assuming a conservative prevalence of 50%, which maximises variance, a sample size of 824 yields a standard error of approximately 0.017 and a 95% confidence interval margin of error of approximately ±3.4 percentage points. The available sample therefore provided good precision for estimating the prevalence of non-spousal primary social supporter selection. For regression analyses, model complexity was considered in relation to the number of observed outcome events to limit the risk of model overfitting.

### Statistical analysis

All analyses were conducted using STATA version 18. Descriptive analyses were conducted to summarise participant characteristics: continuous variables were summarised using median and interquartile range (IQR) or mean and standard deviation (SD), as appropriate, while categorical variables were summarised using frequencies and percentages. Prevalence of non-spousal supporter selection was estimated overall and stratified by key sociodemographic characteristics: maternal age, recruitment site, and marital status because these characteristics were considered particularly relevant to the composition and mobilisation of social support networks. Exact binomial 95% confidence intervals (CI) were calculated.

Associations between explanatory variables and non-spousal supporter selection were assessed using modified Poisson regression with robust standard errors to estimate the prevalence ratio (PR) and 95% CI, as the outcome was not rare and odds ratios from logistic regression would have overestimated the association [30]. Univariable modified Poisson regression was first performed for each explanatory variable. Variables were considered for inclusion in the multivariable model based on conceptual relevance, prior evidence, and statistical association in univariable analyses (p<0.20). Variables considered important a priori were retained irrespective of their univariable p-value. The multivariable model was used to estimate adjusted prevalence ratios (aPRs) and 95% CIs. Statistical significance was assessed using a two-sided p-value <0.05.

Complete-case analysis was performed as missingness across all the variables was minimal (<2% for all variables).

### Ethics statement

This study was conducted in accordance with the Declaration of Helsinki. Ethical approval for the Support Moms study was obtained from the Mbarara University of Science and Technology Research Ethics Committee (MUST-2022-631; Initial approval was granted on 20 October 2022, followed by annual renewals covering the periods 9 September 2023 to 9 September 2024, 1 September 2024 to 1 September 2025, and 9 September 2025 to 9 September 2026) and the Uganda National Council for Science and Technology (HS3366ES). The study participants were enrolled into the Support-Moms study between 27 February and 17 December 2024. Administrative permission was granted by the Mbarara District Health Office, Mbarara City Health Office, Mitooma District Health Office, and the Director of Clinical Services at the Ministry of Health. All participants provided written informed consent prior to enrolment. Pregnant adolescents below 18 years were considered emancipated minors in accordance with applicable Ugandan guidelines and provided written informed consent independently, without requiring parental or guardian consent.

Participation was voluntary, and participants were free to withdraw at any time without consequence. The present manuscript reports a secondary analysis of baseline data collected as part of the Support-Moms study. The dataset was accessed for the purposes of this analysis on 26 January 2026. The authors did not have access to information that could directly identify individual participants.

## Results

### Participants

A total of 824 pregnant women were enrolled in the Support-Moms study, and all were included in this cross-sectional analysis. Complete data were available for the primary outcome, with no exclusions due to missing outcome data. Missingness across covariates was minimal; spouse– woman age difference had 13 missing observations (1.6%), while all other variables were complete.

### Participant characteristics

Over half of the participants were recruited from Mbarara district (56.3%, n=464). Most women were aged 20–34 years (74.9%, n=617), were married/cohabiting (96.6%, n=796), and had attained primary education or less (58.6%, n=483). Nearly three-quarters (73.8%, n=608) were multiparous, and 60.3% (n=497) were enrolled at ≥13 weeks’ gestation. Half of the pregnancies were planned (50.0%, n=412). The median household monthly income was 100,000 UGX (IQR 60,000–240,000), and 47.5% (n=391) of women lived ≥5 km from the nearest health facility (Table 1).

**Table 1:**
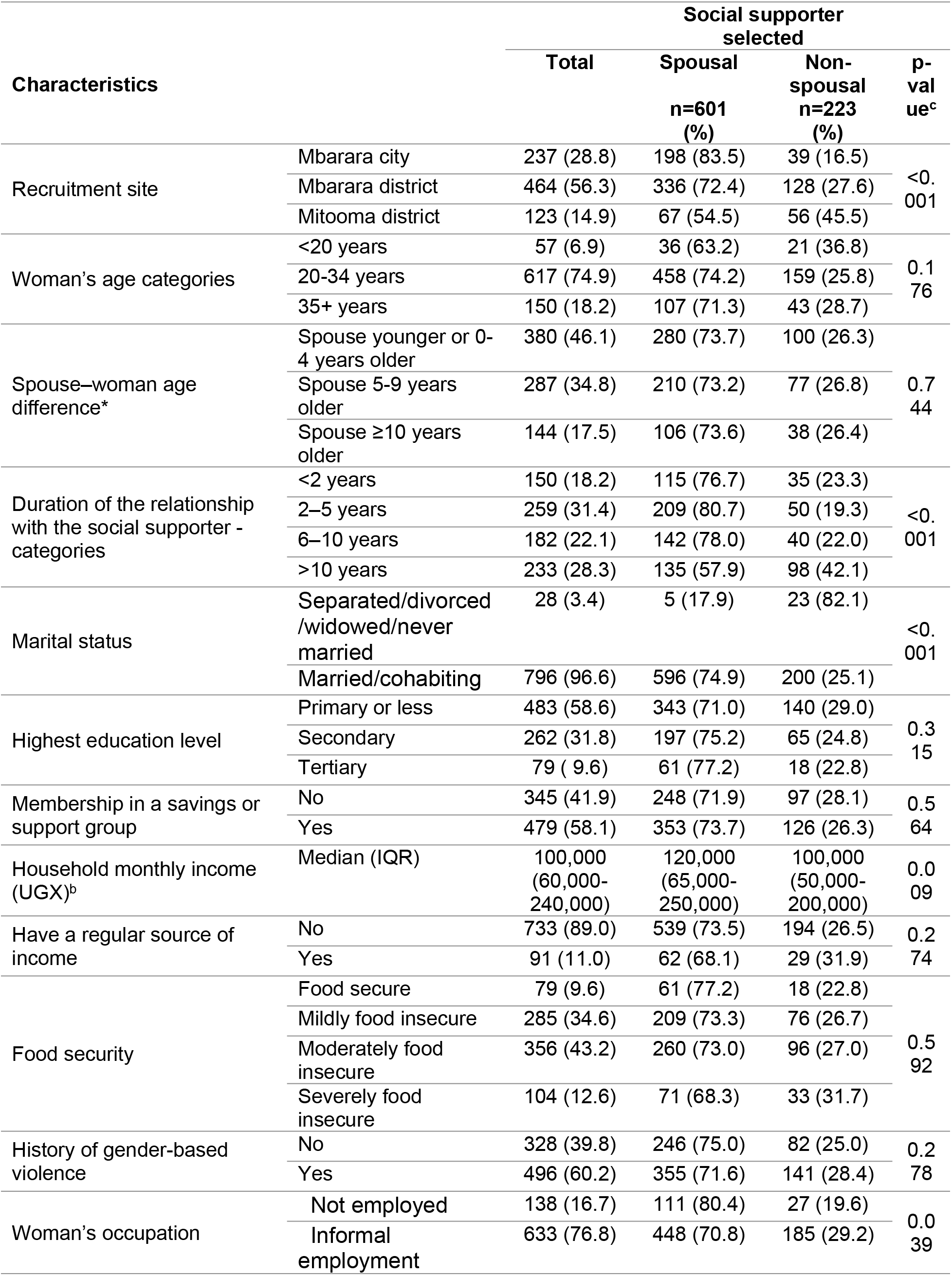
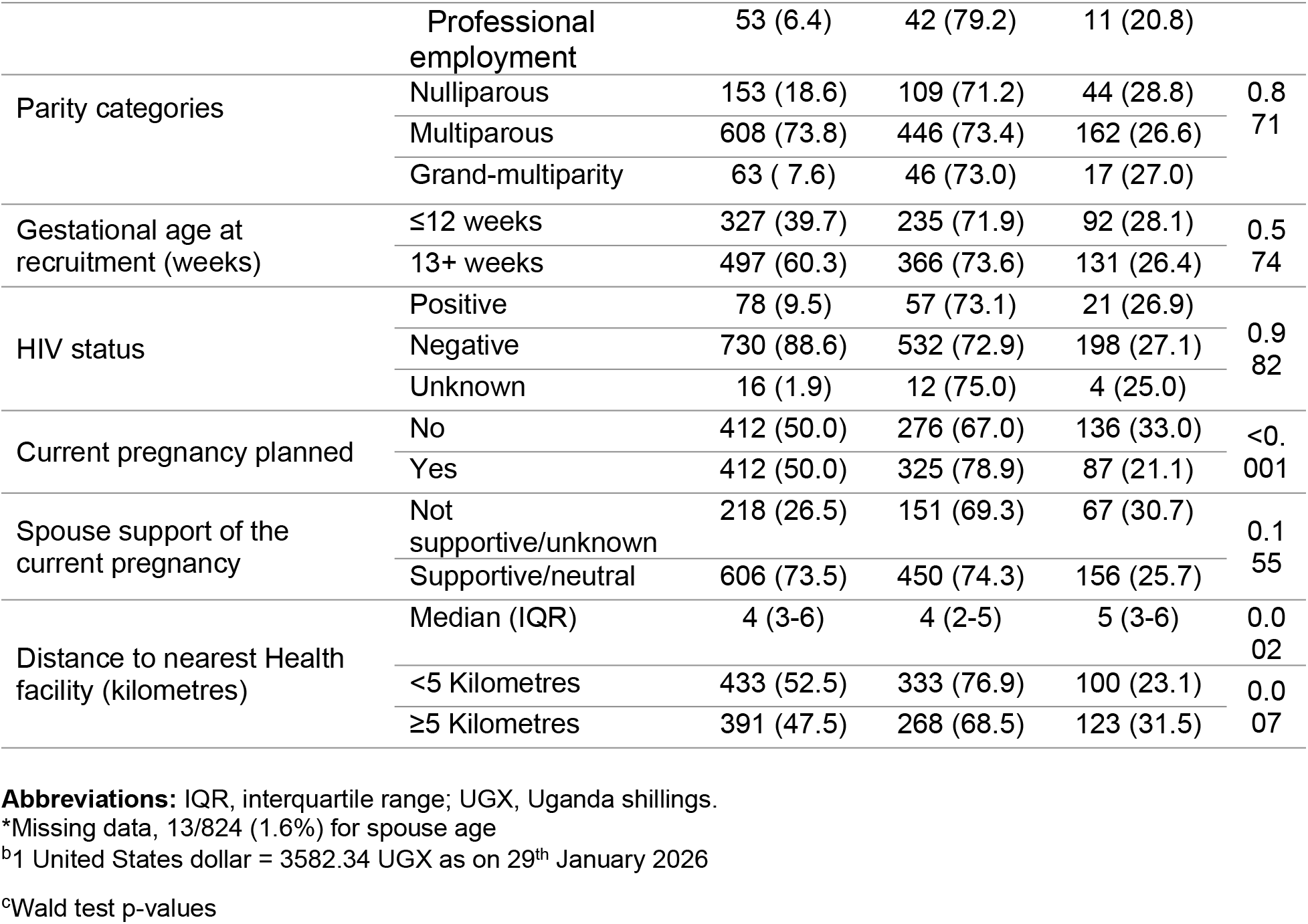
Baseline characteristics of pregnant women at enrolment, overall and stratified by type of social supporter selected, N=824.

| Characteristics |  |  | Social supporter selected |  |  |
|---|---|---|---|---|---|
|  |  | Total | Spousal<br>n=601 (%) | Non-spousal<br>n=223 (%) | p-value<sup>c</sup> |
| Recruitment site | Mbarara city | 237 (28.8) | 198 (83.5) | 39 (16.5) | <0.001 |
|  | Mbarara district | 464 (56.3) | 336 (72.4) | 128 (27.6) |  |
|  | Mitooma district | 123 (14.9) | 67 (54.5) | 56 (45.5) |  |
| Woman's age categories | <20 years | 57 (6.9) | 36 (63.2) | 21 (36.8) | 0.176 |
|  | 20-34 years | 617 (74.9) | 458 (74.2) | 159 (25.8) |  |
|  | 35+ years | 150 (18.2) | 107 (71.3) | 43 (28.7) |  |
| Spouse–woman age difference* | Spouse younger or 0-4 years older | 380 (46.1) | 280 (73.7) | 100 (26.3) | 0.744 |
|  | Spouse 5-9 years older | 287 (34.8) | 210 (73.2) | 77 (26.8) |  |
|  | Spouse ≥10 years older | 144 (17.5) | 106 (73.6) | 38 (26.4) |  |
| Duration of the relationship with the social supporter - categories | <2 years | 150 (18.2) | 115 (76.7) | 35 (23.3) | <0.001 |
|  | 2–5 years | 259 (31.4) | 209 (80.7) | 50 (19.3) |  |
|  | 6–10 years | 182 (22.1) | 142 (78.0) | 40 (22.0) |  |
|  | >10 years | 233 (28.3) | 135 (57.9) | 98 (42.1) |  |
| Marital status | Separated/divorced/widowed/never married | 28 (3.4) | 5 (17.9) | 23 (82.1) | <0.001 |
|  | Married/cohabiting | 796 (96.6) | 596 (74.9) | 200 (25.1) |  |
| Highest education level | Primary or less | 483 (58.6) | 343 (71.0) | 140 (29.0) | 0.315 |
|  | Secondary | 262 (31.8) | 197 (75.2) | 65 (24.8) |  |
|  | Tertiary | 79 ( 9.6) | 61 (77.2) | 18 (22.8) |  |
| Membership in a savings or support group | No | 345 (41.9) | 248 (71.9) | 97 (28.1) | 0.564 |
|  | Yes | 479 (58.1) | 353 (73.7) | 126 (26.3) |  |
| Household monthly income (UGX)<sup>b</sup> | Median (IQR) | 100,000 (60,000-240,000) | 120,000 (65,000-250,000) | 100,000 (50,000-200,000) | 0.009 |
| Have a regular source of income | No | 733 (89.0) | 539 (73.5) | 194 (26.5) | 0.274 |
|  | Yes | 91 (11.0) | 62 (68.1) | 29 (31.9) |  |
| Food security | Food secure | 79 (9.6) | 61 (77.2) | 18 (22.8) | 0.592 |
|  | Mildly food insecure | 285 (34.6) | 209 (73.3) | 76 (26.7) |  |
|  | Moderately food insecure | 356 (43.2) | 260 (73.0) | 96 (27.0) |  |
|  | Severely food insecure | 104 (12.6) | 71 (68.3) | 33 (31.7) |  |
| History of gender-based violence | No | 328 (39.8) | 246 (75.0) | 82 (25.0) | 0.278 |
|  | Yes | 496 (60.2) | 355 (71.6) | 141 (28.4) |  |
| Woman's occupation | Not employed | 138 (16.7) | 111 (80.4) | 27 (19.6) | 0.039 |
|  | Informal employment | 633 (76.8) | 448 (70.8) | 185 (29.2) |  |
|  | Professional employment | 53 (6.4) | 42 (79.2) | 11 (20.8) |  |
| Parity categories | Nulliparous | 153 (18.6) | 109 (71.2) | 44 (28.8) | 0.871 |
|  | Multiparous | 608 (73.8) | 446 (73.4) | 162 (26.6) |  |
|  | Grand-multiparity | 63 ( 7.6) | 46 (73.0) | 17 (27.0) |  |
| Gestational age at recruitment (weeks) | ≤12 weeks | 327 (39.7) | 235 (71.9) | 92 (28.1) | 0.574 |
|  | 13+ weeks | 497 (60.3) | 366 (73.6) | 131 (26.4) |  |
| HIV status | Positive | 78 (9.5) | 57 (73.1) | 21 (26.9) | 0.982 |
|  | Negative | 730 (88.6) | 532 (72.9) | 198 (27.1) |  |
|  | Unknown | 16 (1.9) | 12 (75.0) | 4 (25.0) |  |
| Current pregnancy planned | No | 412 (50.0) | 276 (67.0) | 136 (33.0) | <0.001 |
|  | Yes | 412 (50.0) | 325 (78.9) | 87 (21.1) |  |
| Spouse support of the current pregnancy | Not supportive/unknown | 218 (26.5) | 151 (69.3) | 67 (30.7) | 0.155 |
|  | Supportive/neutral | 606 (73.5) | 450 (74.3) | 156 (25.7) |  |
| Distance to nearest Health facility (kilometres) | Median (IQR) | 4 (3-6) | 4 (2-5) | 5 (3-6) | 0.002 |
|  | <5 Kilometres | 433 (52.5) | 333 (76.9) | 100 (23.1) | 0.007 |
|  | ≥5 Kilometres | 391 (47.5) | 268 (68.5) | 123 (31.5) |  |

**Abbreviations:** IQR, interquartile range; UGX, Uganda shillings.
\*Missing data, 13/824 (1.6%) for spouse age
<sup>b</sup>1 United States dollar = 3582.34 UGX as on 29<sup>th</sup> January 2026
<sup>c</sup>Wald test p-values

Non-spousal supporter selection varied by recruitment site, with the highest prevalence among women recruited in Mitooma District, the predominantly rural study setting, and the lowest among those recruited in Mbarara City, the predominantly urban study setting (45.5% versus 16.5%, p<0.001). Women who were separated, divorced, widowed or never married were substantially more likely to select a non-spousal supporter than those married or cohabiting (82.1% versus 25.1%, p<0.001). Similarly, women reporting a relationship duration greater than 10 years were more likely to select a non-spousal supporter (42.1%) compared to those in shorter relationships. Non-spousal supporter selection was also more common among women with unplanned pregnancies (33.0% versus 21.1% among planned pregnancies) and among those residing ≥5 km from a health facility (31.5% versus 23.1%) (Table 1).

### Prevalence of non-spousal social supporter selection

Overall, 223 of 824 pregnant women selected a non-spousal social supporter at enrolment into the parent study, corresponding to a prevalence of 27.1% (95% CI 24.1–30.3). Prevalence varied across selected participant characteristics (Figure 1). It was highest among women recruited in Mitooma District, the predominantly rural study setting (45.5%, 95% CI 36.5–54.7), and lowest among those recruited in Mbarara City, the predominantly urban study setting (16.5%, 95% CI 12.3–21.8). Women aged <20 years had a prevalence of 36.8% (95% CI 24.4–51.3), compared with 25.8% among those aged 20–34 years and 28.7% among those aged ≥35 years. Marked differences were observed by marital status: 82.1% (95% CI 63.1–93.9) of women who were separated, divorced, widowed, or never married selected a non-spousal supporter, compared with 25.1% (95% CI 22.2–28.3) among women who were married or cohabiting.

**Fig 1:**
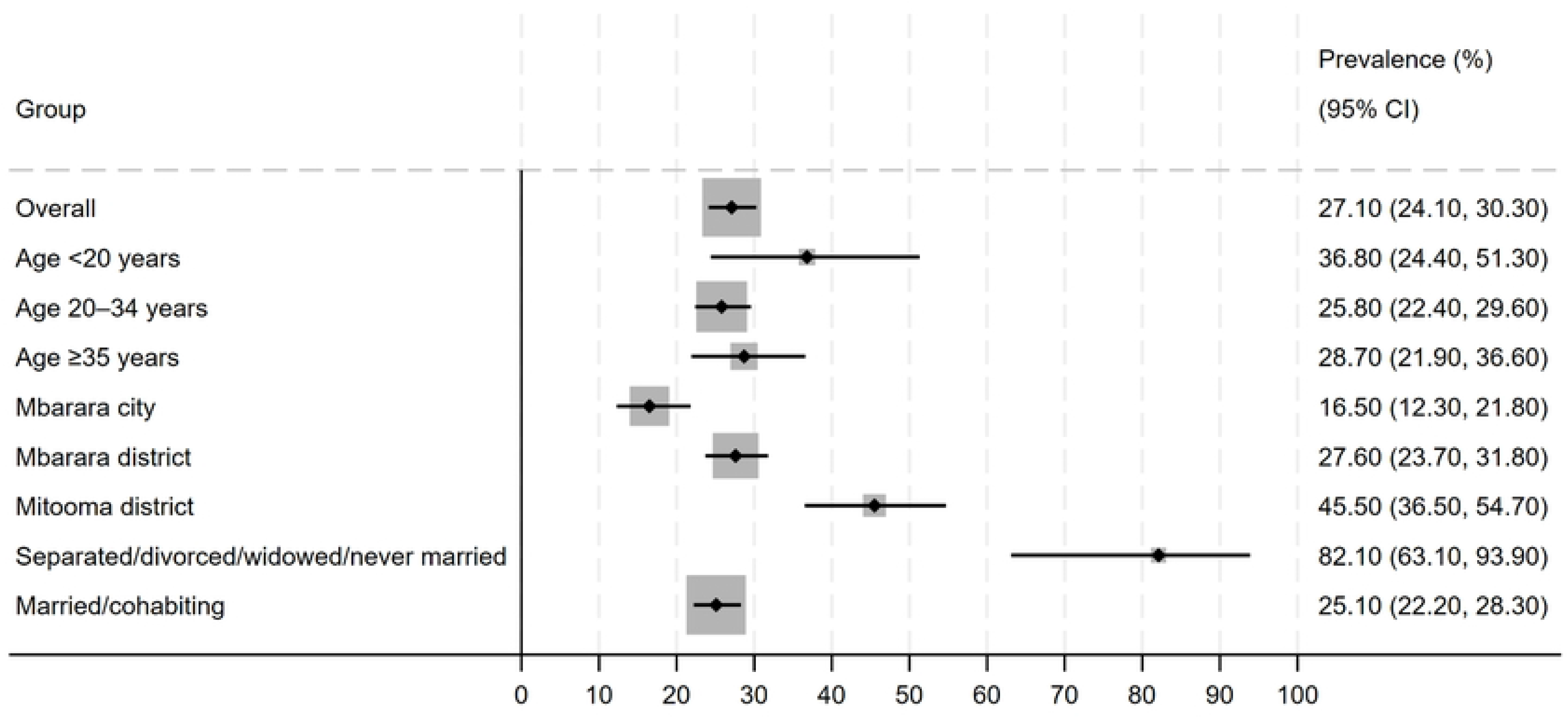
Prevalence of non-spousal social supporter selection among pregnant women in Southwestern Uganda, overall and by selected subgroups. Points represent prevalence estimates, and horizontal bars indicate 95% confidence intervals.

Among the 223 women who selected a non-spousal primary supporter, friends were the most frequently nominated (68/223, 30.5%), followed by other relatives (64/223, 28.7%); together, these groups accounted for 59.2% of non-spousal nominations. Siblings (40/223, 17.9%) and mothers (37/223, 16.6%) accounted for a further 34.5%, while neighbours (8/223, 3.6%) and VHTs/healthcare providers (6/223, 2.7%) were less frequently nominated.

### Univariable analysis

Compared with women recruited in Mbarara City, those recruited in Mbarara District had a 68% higher prevalence of non-spousal primary supporter selection (cPR: 1.68, 95% CI: 1.21–2.31), while those recruited in Mitooma District had nearly three times the prevalence (cPR: 2.77, 95% CI: 1.96–3.91). Women who were separated, divorced, widowed, or never married had more than three times the prevalence of selecting a non-spousal supporter compared to women who were married or cohabiting (cPR: 3.27, 95% CI 2.65–4.03). Women reporting an unplanned pregnancy had a higher prevalence of selecting a non-spousal supporter compared to those with a planned pregnancy (cPR: 1.56, 95% CI: 1.24–1.97). Living ≥5 km from the nearest health facility was also associated with increased prevalence (cPR: 1.36, 95% CI: 1.09–1.71). Women whose relationship with the nominated social supporter had lasted >10 years had more than twice the prevalence of non-spousal supporter selection compared with women whose relationship had lasted 2–5 years (cPR: 2.18, 95% CI: 1.63–2.92). Informal employment was associated with a higher prevalence compared to unemployment (cPR: 1.49, 95% CI: 1.04–2.14). Maternal age, spouse–woman age difference, gestational age at enrolment, parity, education level, household income, food security, history of gender-based violence, spouse support of the pregnancy, HIV status, and savings group membership were not significantly associated with non-spousal supporter selection in univariable analysis (Table 2).

**Table 2:** Univariable analysis for the factors associated with non-spousal social supporter selection among pregnant women, N=824.

| **Characteristics** |  | **Crude PR (95% CI)** | **p-value<sup>c</sup>** |
|---|---|---|---|
| Woman’s age categories | <20 years | 1.43 (0.99-2.06) | 0.055 |
|  | 20-34 years | Reference |  |
|  | 35+ years | 1.11 (0.84-1.48) | 0.465 |
| Spouse–woman age difference* | Spouse younger or 0–4 years older | 0.98 (0.76-1.27) | 0.882 |
|  | Spouse 5-9 years older | 1.00 |  |
|  | Spouse ≥10 years older | 0.98 (0.70-1.37) | 0.922 |
| Gestational age categories (weeks) | ≤12 weeks | Reference | 0.574 |
|  | 13+ weeks | 0.94 (0.75-1.18) |  |
| Duration of the relationship with the social supporter | <2 years | 1.21 (0.82-1.77) | 0.332 |
|  | 2–5 years | Reference |  |
|  | 6–10 years | 1.14 (0.79-1.65) | 0.492 |
|  | >10 years | 2.18 (1.63-2.92) | <0.001* |
| Marital status | Separated/divorced/widowed/never married | 3.27 (2.65-4.03) | <0.001* |
|  | Married/cohabiting | Reference |  |
| Highest education level | Primary or less | 1.27 (0.83-1.95) | 0.272 |
|  | Secondary | 1.09 (0.69-1.72) | 0.715 |
|  | Tertiary | Reference |  |
| Membership in a savings or support group | No | 1.07 (0.85-1.34) | 0.563 |
|  | Yes | Reference |  |
| Household monthly income<sup>b</sup> | ≤150,000 UGX | 1.08 (0.86-1.37) | 0.510 |
|  | >150,000 UGX | 1.00 |  |
| Current pregnancy planned | No | 1.56 (1.24-1.97) | <0.001* |
|  | Yes | Reference |  |
| Spouse support of the current pregnancy | Not supportive/unknown | 1.19 (0.94-1.52) | 0.149 |
|  | Supportive/neutral | Reference |  |
| Distance from nearest health facility | <5 Kilometres | Reference | 0.007* |
|  | ≥5 Kilometres | 1.36 (1.09-1.71) |  |
| Recruitment site | Mbarara city | Reference |  |
|  | Mbarara district | 1.68 (1.21-2.31) | 0.002* |
|  | Mitooma district | 2.77 (1.96-3.91) | <0.001* |
| Food security | Food secure | Reference |  |
|  | Mild food insecurity | 1.17 (0.75-1.83) | 0.493 |
|  | Moderate food insecurity | 1.18 (0.76-1.84) | 0.454 |
|  | Severe food insecurity | 1.39 (0.85-2.28) | 0.189 |
| History of gender-based violence | No | Reference | 0.282 |
|  | Yes | 1.14 (0.90-1.44) |  |
| HIV status | Positive | Reference |  |
|  | Negative | 1.01 (0.69-1.48) | 0.970 |
|  | Unknown | 0.93 (0.37-2.34) | 0.875 |
| Parity categories | Nulliparous | 1.08 (0.81-1.43) | 0.596 |
|  | Multiparous | 1.00 |  |
|  | Grand-multiparity | 1.01 (0.66-1.55) | 0.954 |
| Woman’s occupation | Not employed | 1.00 |  |
|  | Informal employment | 1.49 (1.04-2.14) | 0.029 * |
|  | Professional employment | 1.06 (0.57-1.98) | 0.853 |

**Abbreviations:** CI, confidence interval; PR, prevalence ratio; UGX, Uganda shillings.
\*Missing data, 13/824 (1.6%) for spouse age
<sup>b</sup>1 United States dollar = 3582.34 UGX as on 29<sup>th</sup> January 2026
<sup>c</sup>Wald test p-values

### Multivariable analysis

Compared with women recruited in Mbarara City, those recruited in Mbarara District had a 50% higher prevalence of non-spousal primary supporter selection (aPR: 1.50, 95% CI: 1.09–2.08), while those recruited in Mitooma District had more than twice the prevalence (aPR: 2.52, 95% CI: 1.77–3.60). Women who were separated, divorced, widowed, or never married had more than twice the prevalence of non-spousal supporter selection compared with women who were married or cohabiting (aPR 2.35, 95% CI 1.76–3.14). An unplanned pregnancy remained independently associated with non-spousal supporter selection (aPR: 1.39, 95% CI: 1.10–1.77). A relationship duration >10 years was also associated with nearly twice the prevalence compared with a duration of 2–5 years (aPR: 1.93, 95% CI: 1.39–2.69). Additionally, women aged <20 years had a higher prevalence compared to those aged 20–34 years (aPR: 1.48, 95% CI: 1.01–2.15). Distance from the nearest health facility, occupation, household income, food security status, history of gender-based violence, spouse support of the pregnancy, and HIV status were not independently associated after adjustment (Table 3).

**Table 3:** Adjusted multivariable analysis for the factors associated with non-spousal social supporter selection among pregnant women, N=824.

| Characteristics | Prevalence, n/N (%) | cPR (95% CI) | p-value<sup>b</sup> | aPR (95% CI) | p-value<sup>b</sup> |
|---|---|---|---|---|---|
| **Recruitment site** |  |  |  |  |  |
| Mbarara City | 39/237 (16.5) | Reference |  | 1.00 |  |
| Mbarara District | 128/464 (27.6) | 1.68 (1.21–2.31) | 0.002 | 1.50 (1.09–2.08) | 0.013* |
| Mitooma District | 56/123 (45.5) | 2.77 (1.96–3.91) | <0.001 | 2.52 (1.77–3.60) | <0.001* |
| **Distance to nearest health facility** |  |  |  |  |  |
| <5 kilometres | 100/433 (23.1) | Reference |  | 1.00 | — |
| ≥5 kilometres | 123/391 (31.5) | 1.36 (1.09–1.71) | 0.007 | 1.15 (0.92–1.43) | 0.221 |
| **Current pregnancy planned** |  |  |  |  |  |
| Yes | 87/412 (21.1) | Reference |  | 1.00 |  |
| No | 136/412 (33.0) | 1.56 (1.24–1.97) | <0.001 | 1.39 (1.10–1.77) | 0.007* |
| **Marital status** |  |  |  |  |  |
| Married/cohabiting | 200/796 (25.1) | Reference |  | 1.00 |  |
| Separated/divorced/widowed/never married | 23/28 (82.1) | 3.27 (2.65–4.03) | <0.001 | 2.35 (1.76–3.14) | <0.001* |
| **Duration of relationship with social supporter** |  |  |  |  |  |
| <2 years | 35/150 (23.3) | 1.21 (0.82–1.77) | 0.332 | 1.25 (0.84–1.86) | 0.276 |
| 2–5 years | 50/259 (19.3) | Reference |  | 1.00 |  |
| 6–10 years | 40/182 (22.0) | 1.14 (0.79–1.65) | 0.492 | 1.16 (0.82–1.66) | 0.400 |
| >10 years | 98/233 (42.1) | 2.18 (1.63–2.92) | <0.001 | 1.93 (1.39–2.69) | <0.001* |
| **Maternal age category** |  |  |  |  |  |
| <20 years | 21/57 (36.8) | 1.43 (0.99–2.06) | 0.055 | 1.48 (1.01–2.15) | 0.042* |
| 20–34 years | 159/617 (25.8) | Reference |  | 1.00 |  |
| ≥35 years | 43/150 (28.7) | 1.11 (0.84–1.48) | 0.465 | 0.82 (0.59–1.14) | 0.244 |
| **Spouse support of current pregnancy** |  |  |  |  |  |
| Supportive/neutral | 156/606 (25.7) | Reference |  | 1.00 |  |
| Not supportive/unknown | 67/218 (30.7) | 1.19 (0.94–1.52) | 0.149 | 0.98 (0.76–1.26) | 0.859 |
| **Household monthly income<sup>c</sup>** |  |  |  |  |  |
| >150,000 UGX | 79/307 (25.7) | Reference |  | 1.00 |  |
| ≤150,000 UGX | 144/517 (27.9) | 1.08 (0.86–1.37) | 0.510 | 0.81 (0.64–1.03) | 0.089 |
| **Food security** |  |  |  |  |  |
| Food secure | 18/79 (22.8) | Reference |  | Reference |  |
| Mildly food insecure | 76/285 (26.7) | 1.17 (0.75–1.83) | 0.493 | 0.97 (0.63–1.49) | 0.886 |
| Moderately food insecure | 96/356 (27.0) | 1.18 (0.76–1.84) | 0.454 | 1.09 (0.71–1.67) | 0.689 |
| Severely food insecure | 33/104 (31.7) | 1.39 (0.85–2.28) | 0.189 | 1.11 (0.68–1.80) | 0.678 |
| **History of gender-based violence** |  |  |  |  |  |
| No | 82/328 (25.0) | Reference |  | 1.00 |  |
| Yes | 141/496 (28.4) | 1.14 (0.90–1.44) | 0.282 | 1.10 (0.87–1.40) | 0.412 |
| **Woman's occupation** |  |  |  |  |  |
| Not employed | 27/138 (19.6) | Reference |  | 1.00 |  |
| Informal employment | 185/633 (29.2) | 1.49 (1.04–2.14) | 0.029* | 1.26 (0.89–1.77) | 0.188 |
| Professional employment | 11/53 (20.8) | 1.06 (0.57–1.98) | 0.853 | 1.03 (0.58–1.83) | 0.911 |

**Abbreviations:** aPR, adjusted prevalence ratio; CI, confidence interval; cPR, crude prevalence ratio;  UGX, Uganda shillings.
<sup>b</sup>Wald test p-values

## Discussion

In this analysis of pregnant women in southwestern Uganda, 27.1% nominated a non-spousal social supporter as their primary supporter. Friends and other relatives were the most frequently selected non-spousal supporters, accounting for 30.5% and 28.7% of non-spousal nominations, respectively, while siblings and mothers accounted for a further 17.9% and 16.6%. Non-spousal supporter selection was independently associated with recruitment site, marital status, pregnancy planning, duration of the relationship with the supporter, and younger maternal age.

Male partner involvement has been widely promoted as a cornerstone of antenatal care programming in sub-Saharan Africa, with interventions targeting partner attendance, HIV testing uptake, and shared decision-making during pregnancy [14, 16, 18, 20]. Evidence suggests that male involvement can improve uptake of maternal health services and prevention of mother-to-child transmission of HIV [13, 17]. However, these approaches may not fully capture the diversity of social relationships through which women access pregnancy support. In our study, spouses remained the primary supporters for most women (72.9%), but more than one-quarter (27.1%) nominated someone else as their primary supporter. Among these women, friends and other relatives together accounted for 59.2% of nominations, with mothers and siblings accounting for a further 34.5%. Thus, the finding is not simply one of absent partner support; rather, it demonstrates that a substantial minority of women identified other established social ties as the first person they nominated to support them during pregnancy.

Our findings differ from those of a rural Nigerian mixed-methods study in which 95.2% of women reported expecting support from their husbands and only 4.8% reported not expecting such support [31]. However, these findings are not directly comparable because the Nigerian study assessed expectations of support from husbands, whereas our study assessed the first person women nominated as a social supporter. A woman may therefore expect support from her spouse while simultaneously identifying another person as her primary supporter for the purposes of pregnancy-related support. The higher prevalence of non-spousal primary supporter selection observed in our study may reflect differences in the outcome definition, union status, relationship context, and the social and cultural organisation of support across settings. In addition, the supporter-nomination procedure may have influenced the observed distribution of primary and secondary supporters, because women who did not nominate their spouse first were asked to include their spouse among their secondary supporters.

Several factors were independently associated with non-spousal primary supporter selection. Marital status showed one of the strongest associations: women who were separated, divorced, widowed, or never married had more than twice the prevalence of non-spousal supporter selection compared with women who were married or cohabiting. This finding is consistent with evidence that partner engagement during pregnancy is closely linked to cohabitation and relational stability, and that women without a co-resident spouse may rely more on broader social networks for pregnancy-related support [2, 32–34]. Recruitment site was also associated with supporter selection. Compared with Mbarara City, the predominantly urban study setting, the prevalence was 50% higher among women recruited in Mbarara District and more than twice as high among those recruited in Mitooma District, the predominantly rural study setting. This gradient may reflect differences in social and kinship structures across urban, peri-urban, and rural settings [31], although this cross-sectional analysis cannot establish the mechanisms underlying these differences. Women with unplanned pregnancies also had a higher prevalence of non-spousal supporter selection, consistent with evidence that unintended pregnancy may increase reliance on friends and family for support, possibly reflecting network mobilisation in response to perceived need [23]. Women aged <20 years had a 48% higher prevalence of non-spousal supporter selection than women aged 20–34 years, consistent with literature describing greater reliance on family networks among younger pregnant women [15, 21, 22, 32]. However, our analysis did not establish whether this association was specifically driven by greater selection of mothers or other maternal relatives. Finally, women whose relationship with their nominated supporter had lasted >10 years had a higher prevalence of non-spousal supporter selection than those whose relationship with their nominated supporter had lasted 2–5 years. Because relationship duration was measured for the nominated social supporter, rather than for the spouse or marital relationship, this finding should not be interpreted as evidence that longer marriages increase non-spousal supporter selection. Instead, it may indicate that long-standing social ties can serve as important sources of pregnancy support, although the present analysis cannot determine which supporter types accounted for the longer-duration relationship [21]. History of gender-based violence was not independently associated with non-spousal supporter selection in this analysis. However, the measure captured a history of GBV rather than specifically assessing current intimate partner violence; therefore, the findings cannot establish whether ongoing relationship violence influences women’s choice of primary supporter.

The implications of supporter type for maternal health behaviours remain an important unanswered question. Evidence from sub-Saharan Africa and other low- and middle-income settings suggests that male-partner involvement in antenatal care can improve utilisation of skilled birth attendance and facility delivery [14, 16, 18, 20], supporting continued engagement of spouses where women identify them as important sources of support. At the same time, emerging evidence indicates that other members of women’s social networks can also contribute to maternal healthcare utilisation. Qualitative evidence from Ghana has shown that husbands, in-laws, relatives, and friends may provide informational and instrumental support that facilitates facility delivery [9], while research from Uganda has identified mothers, mothers-in-law, and other female elders as important sources of information about antenatal care initiation [12]. Research from Southwest Ethiopia has similarly reported that support from family members, relatives, and friends/neighbours was associated with health facility delivery [35]. However, direct comparative evidence on whether spousal and non-spousal primary supporters differ in their effectiveness for subsequent antenatal care attendance, facility delivery, or other maternal health behaviours appears limited. This represents an important opportunity for future analyses of the Support-Moms cohort, where the effectiveness of different supporter types can be examined using prospective maternal health outcomes.

### Strengths and limitations

This study has several important strengths. First, it directly examined primary social supporter selection during pregnancy and quantified factors associated with selecting a non-spousal supporter, addressing an understudied aspect of maternal social support. Second, supporter selection was consequential rather than hypothetical: women identified an actual individual from their existing social network who was subsequently engaged as a social supporter within the parent intervention. Thus, the study provides evidence about support relationships that women mobilised in an actual antenatal care context rather than stated preferences in a hypothetical scenario. Third, the analysis considered multiple sociodemographic, reproductive, relationship, and social characteristics simultaneously, allowing estimation of adjusted associations while accounting for potential confounding. Fourth, measuring the duration of the relationship with the nominated supporter provided information about the longevity of social ties beyond marital status alone. Finally, the study was embedded within routine antenatal care and included women recruited across predominantly urban, peri-urban, and rural settings, enhancing the contextual and programmatic relevance of the findings.

Nevertheless, several limitations warrant consideration. First, the cross-sectional design precludes establishing temporality or causality for the observed associations. Second, supporter nomination and several explanatory variables were self-reported and may be subject to recall or social desirability bias, particularly in a context where partner involvement during pregnancy may be socially expected. Third, although the analysis adjusted for several measured characteristics, residual confounding from unmeasured factors such as relationship quality, emotional closeness, or broader social capital remains possible. Fourth, the relationship categories available in the baseline data did not separately distinguish spouses from other intimate partners. Accordingly, the outcome should be interpreted specifically as non-spousal supporter selection rather than selection of a non-intimate partner. Some individuals classified as non-spousal supporters may therefore have been intimate partners or fathers of the pregnancy, but this could not be established from the available data. Fifth, the measure of gender-based violence captured a history of GBV rather than specifically assessing current intimate partner violence. Therefore, the absence of an association with history of GBV should not be interpreted as evidence that ongoing relationship violence does not influence supporter selection. Sixth, the supporter nomination procedure may have influenced the observed distribution of primary and secondary supporters. Participants who did not nominate their spouse as their primary supporter were asked to include their spouse among their secondary supporters; consequently, non-spousal primary selection may partly reflect prioritisation of another social tie within the study’s nomination structure rather than an unconstrained ranking of all available supporters. Finally, the study was conducted in southwestern Uganda and included selected health facilities in Mbarara City, Mbarara District, and Mitooma District; therefore, the findings may not be generalisable to populations with different social, cultural, or health-system contexts.

### Conclusions

More than one-quarter of pregnant women in this study selected a non-spousal primary social supporter, most commonly a friend or other relative. The findings highlight the diversity of social relationships through which women identify support during pregnancy. Maternal health programmes could complement partner-focused approaches by asking women whom they prefer to involve in their care. Further research is needed to determine how different types of support relate to maternal health service engagement and pregnancy outcomes.

## Data Availability

The datasets generated and/or analysed during the current study are not publicly available due to ethical and confidentiality considerations but are available from the corresponding author on reasonable request and with appropriate institutional approvals.

## Declarations

## Acknowledgements

The authors thank the study participants, health facility staff, and the Support Moms research team for their contributions to this work.

## Author contributions

EA, HO, GRM, EBA, JH, PW, VTN, MJS, LT, CL, JA, ST, and LTM contributed to conceptualization of the study. EA, HO, GRM, JH, VN, PW, MK, ST, LTM, and MJS contributed to methodology. HO, EA, MK, and GRM conducted the formal analysis. EA, HO, EBA, JH, GRM, VTN, PW, MK, LT, ST, JA, CL, and CO contributed to investigation and data curation. HO, EA, EBA, JH, VTN, PW, LT, CL, JA, ST, and GRM drafted the manuscript. All authors contributed to interpretation of the findings and critically reviewed and revised the manuscript for important intellectual content. EA, GRM, MJS, CA, and LTM provided supervision, while EA, HO, CA, and GRM contributed to project administration. EA, CA, GRM, MJS, LTM, and PW contributed to funding acquisition and provision of resources. All authors read and approved the final manuscript.

## References

1. Schaffer, M.A. and B. Lia-Hoagberg, Effects of social support on prenatal care and health behaviors of low-income women. J Obstet Gynecol Neonatal Nurs, 1997. 26(4): p. 433–40.

2. Cairney, J., et al., Stress, social support and depression in single and married mothers. Soc Psychiatry Psychiatr Epidemiol, 2003. 38(8): p. 442–9.

3. Mabetha, K., et al., Young women’s social support networks during pregnancy in Soweto, South Africa. Afr J Prim Health Care Fam Med, 2024. 16(1): p. e1–e11.

4. Sah, L.K., E. Hatzidimitriadou, and R.K. Sah, Understanding social support during pregnancy: A qualitative study of women’s lived experiences in Nepal. PLoS One, 2025. 20(10): p. e0333885.

5. Collins, N.L., et al., Social support in pregnancy: psychosocial correlates of birth outcomes and postpartum depression. J Pers Soc Psychol, 1993. 65(6): p. 1243–58.

6. J., F.P., et al., Maternal social support predicts birth weight and fetal growth in human pregnancy. Psychosomatic Medicine, 2000. 62(5): p. 715–725.

7. Stapleton, L.R., et al., Perceived partner support in pregnancy predicts lower maternal and infant distress. J Fam Psychol, 2012. 26(3): p. 453–63.

8. World Health Organization, WHO recommendations on antenatal care for a positive pregnancy experience. 2016: 20 Avenue Appia, 1211 Geneva 27, Switzerland

9. Cofie, L.E., et al., A qualitative study of women’s network social support and facility delivery in rural Ghana. PLoS One, 2018. 13(11): p. e0206429.

10. Berkman, L.F. and T. Glass, Social Integration, Social Networks, Social Support, and Health, in Social Epidemiology, L.F.P.D. Berkman and I.M.D.P.D. Kawachi, Editors. 2000, Oxford University Press. p. 0.

11. Cohen, S. and T.A. Wills, Stress, social support, and the buffering hypothesis. Psychol Bull, 1985. 98(2): p. 310–57.

12. Comfort, A.B., et al., The role of informational support from women’s social networks on antenatal care initiation: qualitative evidence from pregnant women in Uganda. BMC Pregnancy Childbirth, 2022. 22(1): p. 708.

13. Aluisio, A., et al., Male antenatal attendance and HIV testing are associated with decreased infant HIV infection and increased HIV-free survival. J Acquir Immune Defic Syndr, 2011. 56(1): p. 76–82.

14. Byamugisha, R., et al., Determinants of male involvement in the prevention of mother-to-child transmission of HIV programme in Eastern Uganda: a cross-sectional survey. Reproductive Health, 2010. 7(1): p. 12.

15. Engelbrecht, M., N. Mulu, and G. Kigozi-Male, Sesotho Women’s Preferences for Male Partner Involvement During Antenatal Care and Delivery. Int J Environ Res Public Health, 2025. 22(12).

16. Tweheyo, R., et al., Male partner attendance of skilled antenatal care in peri-urban Gulu district, Northern Uganda. BMC Pregnancy and Childbirth, 2010. 10(1): p. 53.

17. Yargawa, J. and J. Leonardi-Bee, Male involvement and maternal health outcomes: systematic review and meta-analysis. J Epidemiol Community Health, 2015. 69(6): p. 604–12.

18. Tokhi, M., et al., Involving men to improve maternal and newborn health: A systematic review of the effectiveness of interventions. PLoS One, 2018. 13(1): p. e0191620.

19. August, F., et al., Community health workers can improve male involvement in maternal health: evidence from rural Tanzania. Glob Health Action, 2016. 9: p. 30064.

20. World Health Organization, WHO recommendations on health promotion interventions for maternal and newborn health 2015. 2015: 20 Avenue Appia, 1211 Geneva 27, Switzerland.

21. Bunting, L. and C. McAuley, Research Review: Teenage pregnancy and motherhood the contribution of support. Child and Family Social Work, 2004. 9(2): p. 207–215.

22. Kotoh, A.M., et al., Pregnant adolescents’ lived experiences and coping strategies in peri-urban district in Southern Ghana. BMC Public Health, 2022. 22(1).

23. Newmyer, L., Who receives support during pregnancy? Variation by intendedness. Soc Sci Res, 2024. 123: p. 103065.

24. Atukunda, E.C., et al., Integration of a Patient-Centered mHealth Intervention (Support-Moms) Into Routine Antenatal Care to Improve Maternal Health Among Pregnant Women in Southwestern Uganda: Protocol for a Randomized Controlled Trial. JMIR Res Protoc, 2025. 14: p. e67049.

25. Atukunda, E.C., et al., Factors Associated with Pregnancy Intentions Amongst Postpartum Women Living with HIV in Rural Southwestern Uganda. AIDS Behav, 2019. 23(6): p. 1552–1560.

26. Atukunda, E.C., et al., Provision of family planning vouchers and early initiation of postpartum contraceptive use among women living with HIV in southwestern Uganda: A randomized controlled trial. PLoS Med, 2019. 16(6): p. e1002832.

27. Young, C.R., et al., Prevalence and correlates of physical and sexual intimate partner violence among women living with HIV in Uganda. PLoS One, 2018. 13(8): p. e0202992.

28. Salvador Castell, G., et al., Household food insecurity access scale (HFIAS). Nutr Hosp, 2015. 31 **Suppl 3**: p. 272–8.

29. Harris, P.A., et al., Research electronic data capture (REDCap)--a metadata-driven methodology and workflow process for providing translational research informatics support. J Biomed Inform, 2009. 42(2): p. 377–81.

30. Barros, A.J.D. and V.N. Hirakata, Alternatives for logistic regression in cross-sectional studies: an empirical comparison of models that directly estimate the prevalence ratio. BMC Medical Research Methodology, 2003. 3(1): p. 21.

31. Arisukwu, O., et al., Spousal Support during Pregnancy in the Nigerian Rural Context. 2020.

32. Figueiredo, B., et al., Teenage pregnancy, attachment style, and depression: a comparison of teenage and adult pregnant women in a Portuguese series. Attach Hum Dev, 2006. 8(2): p. 123–38.

33. Msipu Phiri, T., P. Nyamaruze, and O. Akintola, Perspectives about social support among unmarried pregnant university students in South Africa. PLoS One, 2023. 18(4): p. e0284906.

34. Cohen, K., et al., Partner Involvement During Pregnancy and Maternal Health Behaviors. Matern Child Health J, 2016. 20(11): p. 2291–2298.

35. Mamo, A., et al., Maternal social support and health facility delivery in Southwest Ethiopia. Arch Public Health, 2022. 80(1): p. 135.

